# Multicentric Evaluation of a Novel Point of Care Electrochemical ELISA Platform for SARS-CoV-2 Specific IgG and IgM Antibody Assay

**DOI:** 10.1101/2021.05.04.21256472

**Authors:** Vinay Kumar, Kanad Ghosh, Anagha Chandran, Sachin Panwar, Ananthram Bhat, Shreenivas Konaje, Saroj Das, S Srikanta, Latha Jaganathan, Sujay Prasad, D B Venkatesh, C. Shivaram, P R Krishnaswamy, Navakanta Bhat

**Author notes:** **Corresponding Author :** Vinay Kumar.

## Abstract

New diagnostics technologies for the efficient detection and quantification of SARS-CoV-2 Antibodies is very crucial to manage the COVID-19 pandemic, especially in the context of emerging vaccination paradigms. Herein, we report on a novel point-of-care Electrochemical ELISA platform with disposable screen printed electrodes functionalized with SARS-CoV-2 Spike Glycoprotein S1, to enable fast and accurate quantitative estimation of total antibody concentration (IgG and IgM) in clinical samples. The quantification is performed with a comparison of electrochemical redox current against the current produced by the spiked monoclonal antibodies with known concentration. The assay is validated through multicentric evaluation against 3 different FDA authorized Laboratory standard techniques, using both EDTA whole blood and serum samples. We demonstrate that the proposed assay has excellent sensitivity and specificity, making it a suitable candidate for epidemiological surveys and quantification of antibodies in COVID-19 vaccination programs.

## 1. Introduction

The COVID-19 pandemic, resulting from the SARS-CoV-2 coronavirus has impacted the entire world in an unprecedented manner. Since its identification in December 2019, the pandemic has affected more than 130 million people worldwide, causing 2.9 million fatalities. With the second wave of the disease crippling the world currently, newer and efficient diagnostics technologies become essential in managing the disease. In this context, serological tests become extremely important, which detect SARS-CoV-2 specific antibodies (IgG and IgM), produced typically after the first week of infection, as an immune response from the body [1-3]. The serological tests have also gained a lot of importance recently, especially to assess the efficacy of vaccination towards herd immunity, given the mass vaccination drives launched in several countries. For such applications, there is a need for accurate and reliable point-of-care ELISA platforms for the quantitative measurement of SARS-CoV-2 antibodies.

Since the introduction of serology tests reported in early 2020, there has been a lot of progress in different technologies used for the detection of SARS-CoV-2 specific IgG/IgM antibodies. Almost all the assays primarily target either Spike protein (S1/S2), Nucleoprotein (N) or Specific Receptor Binding Domains (RBD) of SARS-CoV-2, thus enabling specificity [4]. Many studies have been reported in the literature to compare the efficacies of various COVID-19 antibody kits [5-7]. The commercial point-of-care serological tests are primarily based on lateral flow assays [8]. While these are convenient, they are qualitative at best, and require manual intervention for the interpretation of colorimetry results. Liu and Rusling [9] have recently presented a comprehensive review of different technologies used in serology tests. Most of the commercial serological tests till date are predominantly based on optical detection, although there have been some recent attempts to develop electrochemical assays for SARS-CoV-2 serological tests [10,11]. Mahshid, et. al. [10] have highlighted the need for point-of-care SARS-CoV-2 serology tests, especially given the success of electrochemical glucose sensors over the last several decades. If the mature electrochemical sensing technology of Glucometers based on screen printed electrodes can be repurposed, then it could potentially offer an accurate and low-cost solution, for SARS-CoV-2 serology tests. Yakoh et. al. [11] have attempted to show a proof of concept lab scale paper based electrochemical sensors for SARS-CoV-2 antibodies and antigens on a very small set of 17 samples. Furthermore their technique required very elaborate surface functionalization steps, sample preparation steps and testing procedure, making it impractical for point-of-care applications.

In this work we repurpose an existing “Lab on Palm”, electrochemical sensing platform [12-14] and adapt it for the quantitative measurement of SARS-CoV-2 total antibody (IgG/IgM) measurement in clinical whole blood and serum samples. The technology makes use of standard screen printed disposable test strips, with simple surface functionalization process for SARS-CoV-2 specific immunoreceptor, making it amenable for mass manufacturing and deployment. This novel assay has been extensively validated through multicentric evaluation at 4 centre. The assay achieves 100% sensitivity and specificity as compared to 3 different FDA authorized Laboratory standard techniques namely Siemens COV2T S1RBD assay, DiaSorin LIAISON® SARS-CoV-2 S1/S2 IgG assay and Vitros CoV2G IgG assay.

## 2. Materials and Methods

All the SARS-CoV-2 related reagents were procured from The Native Antigen Company, UK. This includes the immunoreceptor used in the assay which is SARS-CoV-2 Spike Glycoprotein (S1) terminally tagged with a predominantly monomeric Sheep Fc-Tag (produced in HEK293 cells) and subsequently conjugated with electrochemically active Horseradish Peroxidase (HRP); Antibodies for spiking experiments, namely the Human recombinant monoclonal IgM Anti-SARS-CoV-2 Spike (S1) Antibody and Human recombinant monoclonal IgG1 Anti-SARS-CoV-2 Spike (S1) Antibody. In addition Goat anti Human IgG from the same vendor was also used for non-binding assay control experiments. The standard bare carbon screen printed electrodes were contract manufactured by GSI Technologies, USA as per the designs provided by PathShodh Healthcare Pvt. Ltd. All other chemicals were procured from Sigma Aldrich (Merck), mainly Phosphate Buffered Saline (PBS), Normal Saline (NaCl) and Stabilcoat Immunoassay stabilizer.

About 6μL solution of immunoreceptor with 120 nM concentration of S1 spike Glycoprotein was prepared in PBS and Satbilcoat and dispensed on working electrode of the carbon printed test strip, using an automated dispensing equipment (BioDot), thereby ensuring manufacturability. The electrodes were dried for an hour at room temperature and a cellulose membrane was taped on the electrodes to serve as protection layer as well as test sample spreading layer during testing. The test strips were packaged in set of 25 tests in dry vials with humidity absorber.

The electrochemical measurement was performed with PathShodh’s multi-analyte diagnostic platform anuPath™. This platform provides a very versatile programmability for a variety of electrochemical techniques. In particular, for the SARS-CoV-2 ELISA, it is repurposed to carry out highly sensitive square wave voltammetry (SWV), which eliminates capacitive noise currents, thus enabling accurate analyte detection. This platform is also capable of storing more than 50,000 patient records and transfer data to any Bluetooth enabled device.

The clinical samples, EDTA whole blood and serum, used in this study were collected from 4 different centres, namely Samatvam Endocrinology Diabetes Centre, Bangalore, Medical Services Trust, Bangalore, Neuberg Anand Diagnostic Laboratory, Bangalore and Manipal Hospital, Bangalore from subjects presenting themselves for COVID-19 antibody testing. All necessary institutional ethical clearances and consents were obtained. In addition, the clinical validation and test licence (MD-13) was obtained from the Indian regulatory body, Central Drugs Standard Control Organisation (CDSCO). The samples tested in one of the laboratory analyzers, were also tested using electrochemical ELISA on the same day, but no later than one week’s time. Serum samples were stored in deep freezer, when they had to be tested beyond the day of the collection, while EDTA whole blood samples were used within 24 hours of collection. Serum samples were allowed to thaw and stabilize to room temperature just before performing the electrochemical ELISA test.

The electrochemical ELISA test is performed by mixing 10 μL of sample (EDTA whole blood or serum) is mixed with 40 μL of 500 mM NaCl buffer. The resultant 50 μL sample is dispensed on the active area (working and counter electrodes). After 4 minutes of waiting time, the square wave voltammetry is performed and the peak current compared against the reference values to first assess if the sample is positive or negative for SARS-CoV-2 specific antibodies and if it is positive, then estimate the concentration.

## 3. Results and Discussions

We use the SARS-CoV-2 S1 Spike Glycoprotein conjugated with Horse Radish Peroxidase as the electrochemically active immunoreceptor (Antigen) in this ELISA assay. A very small volume (6 μL) of 120 nM concentration of immunoreceptor, dissolved in phosphate buffer saline/stabilcoat solution is dispensed on the printed carbon working electrodes of the disposable test strips (Fig. 1a). The hand held electrochemical workstation, anuPath™ (Fig. 1b) is programmed to perform highly sensitive square wave voltammetry measurement to obtain the faradaic redox current from the immunoreceptor. Fig. 1c illustrates the typical reduction current peak obtained from the Antigen in NaCl buffer. When the buffer is spiked with 300nM of SARS-CoV-2 specific antibody (IgG and IgM), the reduction current peak decreases as shown in Fig. 1c. This decrease is due to the fact that the specific antigen-antibody binding results in lower concentration of free antigen available for producing the redox current. Thus the change in reduction current is directly proportional to the amount of antibody present in the test solution.

**Fig .1.**
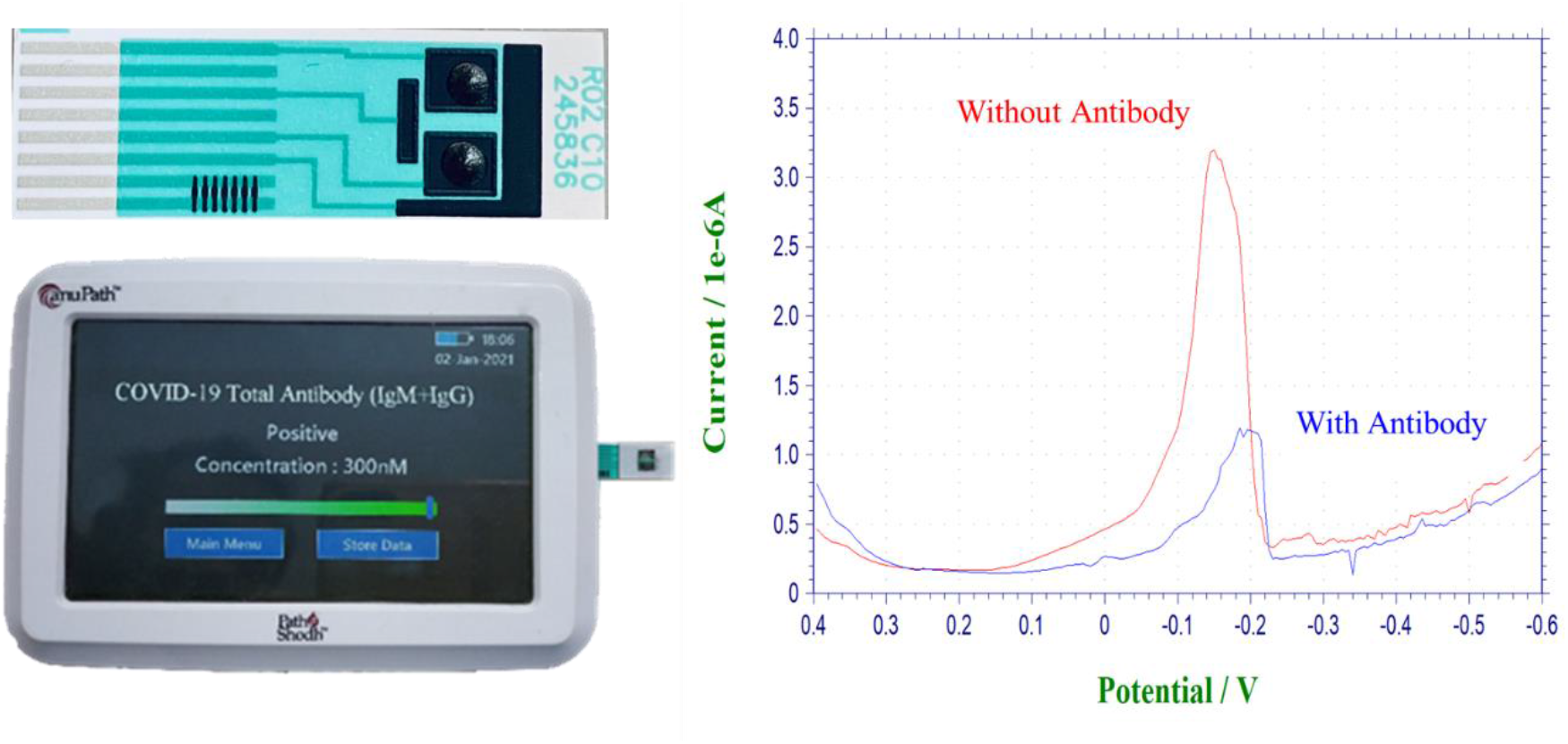
The carbon printed disposable test strip with two working electrodes, 1 counter and 1 reference electrode; Handheld ELISA analyzer; Representative reduction current response in buffer solution without antibody and with antibody.

In order to arrive at the quantitative estimation of antibody concentration, the change in reduction current needs to be calibrated against the known antibody concentration. This was achieved by taking known negative EDTA whole blood and Serum samples from different centres and spiked with known concentration of SARS-CoV-2 monoclonal IgG and IgM antibodies. Fig. 2 shows the calibration curve obtained from the functional relation between the change in current versus spiked antibody concentration in whole blood. Each data point was obtained from triplicate measurement of sample on 3 different test strips, to assess comprehend any statistical variation. It is interesting to note that there are two distinct regions, namely 0-50 nM antibody concentration where the current decreases rapidly, 50-300 nM antibody concentration where the current decrease is rather less. This is because the change in current depends on two factors, namely the concentration of antibodies and the availability of free antigen, from the total copies of antigen on the test strip (4.3×10^11^). When the concentration of antibodies is very low, the availability of antigen for binding is not a rate limiting step and hence the current decreases linearly with increasing concentration of antibodies at a rapid rate. The 50nM antibody concentration in 10 μL sample volume corresponds to 3×10^11^ copies of antigen, which becomes comparable to total antigen concentration. This impacts the availability of free antigen which becomes rate limiting step, resulting in much slower decrease in current. Further both IgG and IgM show similar trends as the assay current depends on free antigen concentration available after the antibody binding process.

**Fig. 2.**
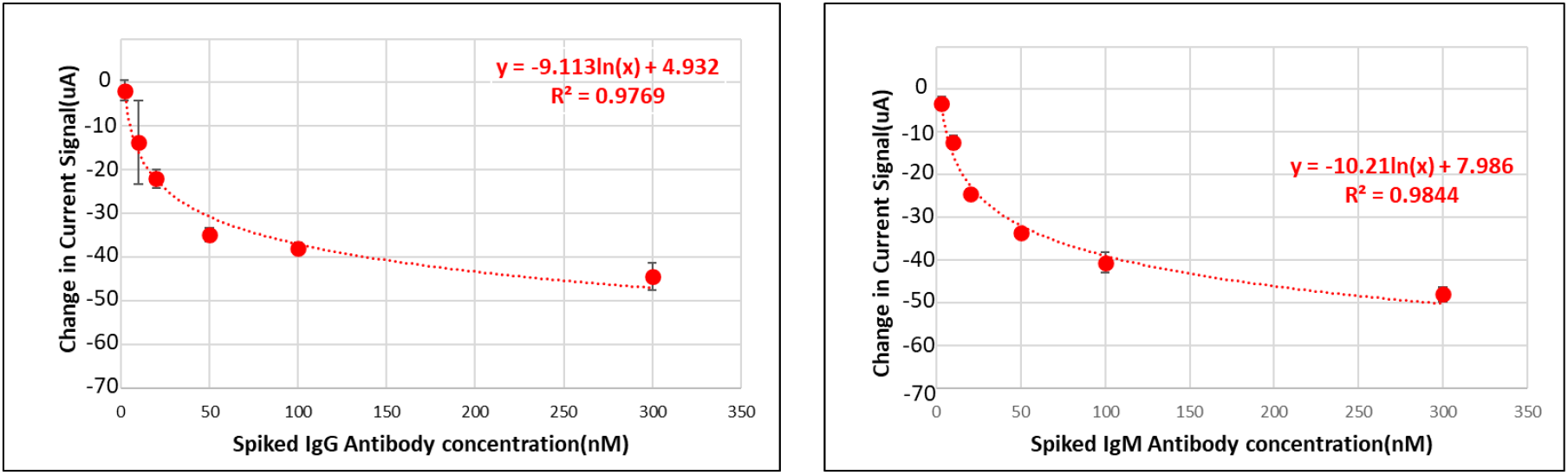
Change in peak reduction current in EDTA whole blood samples as a function of Spiked IgG & IgM concentration

Similarly, Fig. 3 shows the calibration curve obtained from the functional relation between the change in current versus spiked antibody concentration in known negative serum samples. The current response in serum is similar to the EDTA whole blood, indicating the suitability of proposed technique for both whole blood and sera sample evaluation.

**Fig 3.**
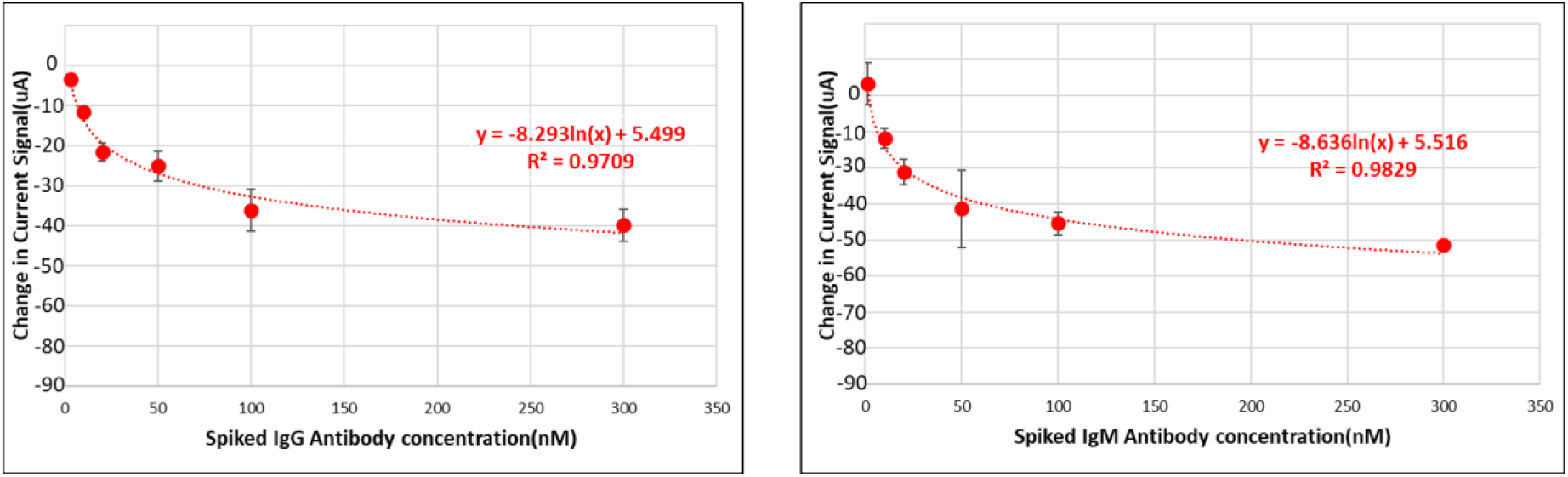
Change in peak reduction current in serum samples as a function of Spiked IgG & IgM concentration

With this initial evaluation and the development of quantification algorithm, large scale validation experiments were conducted at 4 centres, over a period of 4 months. Appropriate ethical clearance and consent were obtained for this study. There was no specific recruitment for the purpose of this study, instead the samples were collected from the subjects, presenting themselves to the participating centre for their routine evaluations and/or getting COVID-19 serology tests done. The EDTA whole blood and/or serum samples were collected at the centres and the SARS-CoV-2 specific antibodies were tested using one of the lab analyzers, described in Materials and Methods section. The same samples were also tested on 5 different anuPath™ analyzers. In order to account for any statistical variation across different analyzers, the samples were randomly assigned to 5 different analyzers for testing. The typical testing protocol included, pipetting 10 μL of sample in a clean cuvette, wherein 40 μL of NaCl buffer solution was added to dilute the sample. The 50 μL sample was dispensed on the test strip that was inserted into the testing port of the analyzer. The sample was allowed to react with the sensing chemistry for a duration of 4 minutes. Then the square wave voltammetry was performed on the analyzer to measure the peak reduction current and detect whether the sample is positive/negative for SARS-CoV-2 specific antibodies. In order to ensure that the assay achieves 100% specificity, the threshold current was set to a value corresponding to 20nM antibody concentration. Table 1 summarizes the results from this study. A total of 460 samples were evaluated, of which 252 were EDTA whole blood samples and 198 were sera samples. The anuPath™ electrochemical ELISA assay achieved excellent correlation with FDA authorized lab analyzers, with sensitivity and specificity of 100%.

**Table 1.** Performance evaluation of anuPath™ Electrochemical ELISA Analyzer

|  | EDTA Whole Blood Samples |  | Sera Samples |  |
| --- | --- | --- | --- | --- |
|  | Lab Analyzer | anuPath™ | Lab analyzer | anuPath™ |
| Negative samples | 120 | 120 | 110 | 110 |
| Positive samples | 132 | 132 | 88 | 88 |
| anuPath™ Sensitivity |  | 100% |  | 100% |
| anuPath™ Specificity |  | 100% |  | 100% |

Fig. 4 demonstrates the correlation between the quantitative value of total antibody concentration expressed in nano molar units, (estimated from the calibration plots in Fig. 2 and 3) and the optical density value in AU/mL measured by the lab analyzers. For both EDTA whole blood and sera samples, we observe an R^2^ value of greater than 0.85, indicating good performance from the point-of-care electrochemical ELISA analyzer.

**Fig. 4.**
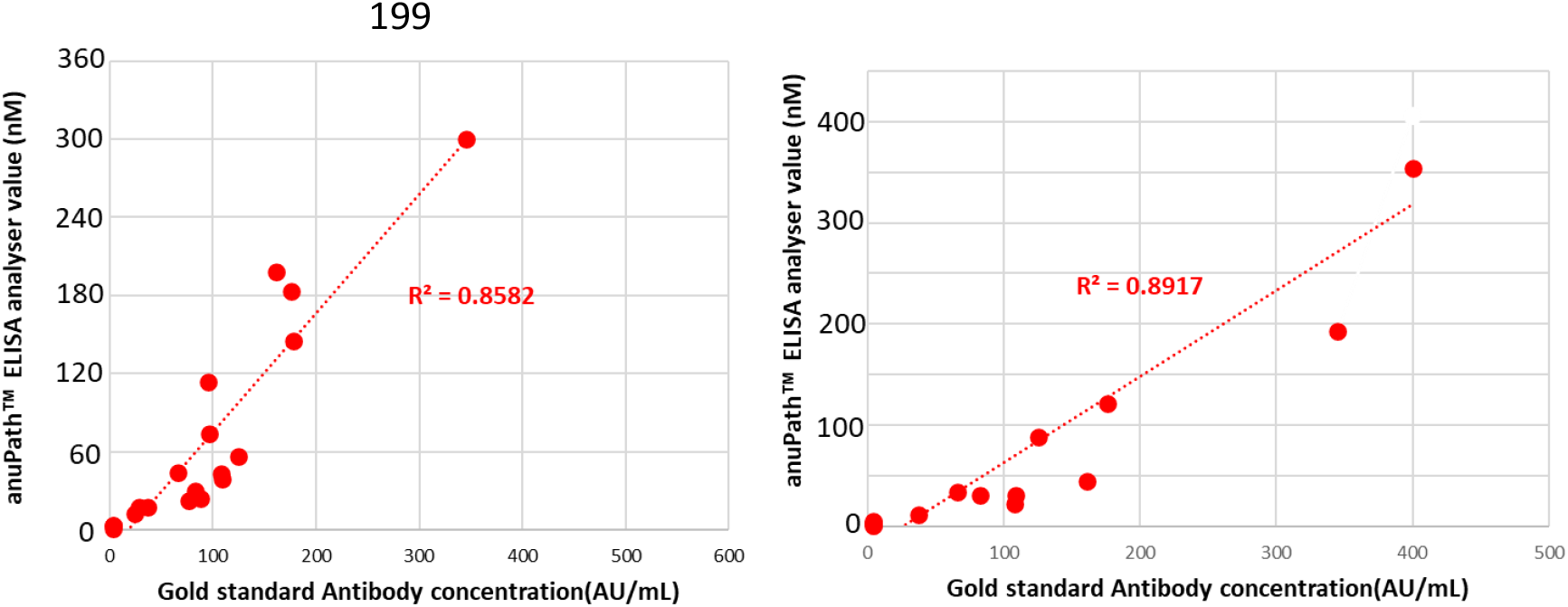
Correlation between anuPath™ electrochemical analyzer and lab analyzer in quantification of SARS-CoV-2 antibody concentration

This electrochemical assay has now been validated independently by Translational Health Science and Technology Institute (THSTI), Faridabad, and is found to meet the regulatory specifications prescribed by the Indian Council for Medical Research (ICMR).

## 4. Conclusions

We demonstrate the efficacy of a novel point-of-care electrochemical ELISA assay through multi-centric evaluation for the detection and quantification of SARS-CoV-2 specific antibodies. The assay uses carbon screen printed electrodes on a disposable test strip, functionalized with SARS-CoV-2 Spike Glycoprotein (S1) C terminally tagged with a Sheep Fc-Tag and conjugated with electrochemically active Horseradish Peroxidase (HRP). The handheld ELISA analyzer implements highly sensitive square wave voltammetry to measure the reduction current peak from the antigen, which is inversely proportional to the concentration of SARS-CoV-2 specific IgG and IgM antibody concentration. The assay has been validated very comprehensively using 460 samples of EDTA whole blood and sera. The proposed assay achieves excellent specificity and sensitivity as compared with FDA authrorized lab analyzers. Given the capabilities such as quantification of SARS-CoV-2 specific antibodies, ease of use, portability, we expect this technology to be very useful in serological surveys and evaluation of COVID-19 vaccine efficacy.

## Data Availability

All the data reported in the manuscript is available for verification

## Credit authorship contribution statement

NB, VK conceived the technology and planned the detailed experimental strategy. PRK, SS, LJ, SP, DBV, CS coordinated the multicentre trials along with the management of samples and evaluation on FDA authorized lab analyzers. VK coordinated all the experiments on developing the electrochemical assay, with KG, AC, SP, AB, SK and SD performing all the experiments with ELISA analyzer. NB wrote the first draft of the manuscript with help from VK. All authors participated in the review and approval of final manuscript.

## Acknowledgements

We would like to thank Dr. Renu Swarup, Dr Kalaivani Ganesan and Dr Kamakshi Chaithri from the Department of Biotechnology, Government of India for facilitating regulatory validation process. We thank Prof. Guruprasad Medigeshi, Anatharaj A, and the Biorepository at the Translational Health Science and Technology Institute (THSTI), Faridabad for conducting the independent validation of this technology. We also thank Indian Council for Medical Research (ICMR) and Central Drugs Standard Control Organisation (CDSCO) for streamlined regulatory processes.

## References

[1] F. Krammer, V. Simon, Serology assays to manage COVID-19, Science. 368 (2020) 1060.

[2] A.K. Winter, S.T. Hegde, The important role of serology for COVID-19 control, Lancet Infect. Dis. 20 (2020) 758–759.

[3] Q.X. Long, et. al., Antibody responses to SARS-CoV-2 in patients with COVID-19, Nat. Med. 26 (2020) 845–848.

[4] Mario Poljak, Anja Ostrbenk Valencak, Tina Stamol, Katja Seme, Head-to-head comparison of two rapid high-throughput automated electrochemiluminescence immunoassays targeting total antibodies to the SARS-CoV-2 nucleoprotein and spike protein receptor binding domain, J. Clin. Virol. 137 (2021) 104784

[5] Michael C. Wehrhahn, et. al., An evaluation of 4 commercial assays for the detection of SARS-CoV-2 antibodies in a predominantly mildly symptomatic low prevalence Australian population, J. Clin. Virol. 138 (2021) 104797

[6] Alix T. Coste, et. al., Comparison of SARS-CoV-2 serological tests with different antigen targets, J. of Clin. Virol. 134 (2021) 104690

[7] Susmita Chaudhuri, et. al., Comparative evaluation of SARS-CoV-2 IgG assays in India, J. of Clin. Virol. 131 (2020) 104609

[8] Steven E. Conklin, et. al., Evaluation of Serological SARS-CoV-2 Lateral Flow Assays for Rapid Point-of-Care Testing, J. Clin. Microbiol. (2021) e02020–20

[9] Guoqiang Liu and James F. Rusling, COVID-19 Antibody Tests and Their Limitations, ACS Sens. 6 (2021) 593−612.

[10] Sahar Sadat Mahshid, et. al., The potential application of electrochemical biosensors in the COVID-19 pandemic: A perspective on the rapid diagnostics of SARS-CoV-2, Biosensors and Bioelectronics 176 (2021) 112905

[11] Abdulhadee Yakoh, et. al., Paper-based electrochemical biosensor for diagnosing COVID-19: Detection of SARS-CoV-2 antibodies and antigen, Biosensors and Bioelectronics 176 (2021) 112912

[12] Vinay Kumar, et. al., Application of a nanotechnology-based, point-of-care diagnostic device in diabetic kidney disease, Kidney international reports. 3 (2018) 1110-1118 (2018)

[13] Vinay Kumar, et. al., Creatinine-Iron Complex and its Use in Electrochemical Measurement of Urine Creatinine, IEEE Sensors Journal 18 (2018) 830–836

[14] Vinay Kumar, et. al., Aza-heterocyclic Receptors for Direct Electron Transfer Hemoglobin Biosensor” Nature Scientific Reports. 7 (2017) 42031

[15] https://www.siemens-healthineers.com/en-in/laboratory-diagnostics/assays-by-diseases-conditions/infectious-disease-assays/cov2t-assay

[16] https://www.diasorin.com/en/immunodiagnostic-solutions/clinical-areas/infectious-diseases/covid-19

[17] https://www.fda.gov/media/137363/download

